# Trends of long-term opioid therapy and subsequent discontinuation among people with chronic non-cancer pain in UK primary care: a retrospective cohort study

**DOI:** 10.1101/2025.02.02.25321533

**Authors:** Qian Cai, Yun-Ting (Joyce) Huang, Thomas Allen, Charlotte Morris, Christos Grigoroglou, Evangelos Kontopantelis

## Abstract

**Objectives:** To examine trends of opioid use, focusing on long-term opioid therapy (L-TOT) and its discontinuation among people with chronic non-cancer pain (CNCP).

**Design and setting:** Retrospective cohort study using UK Clinical Practice Research Datalink Aurum data.

**Population:** Incident opioid users (no opioid use in the prior year) with CNCP between 01/01/2009-31/12/2019. Among them, we identified L-TOT users (≥3 opioid prescriptions within 90 days, or total ≥90 supply days within the first year, excluding the initial 30 days) and L-TOT discontinuers (no opioid use for ≥180 days following a L-TOT).

**Main outcome measures:** Yearly rates of incident opioid users (over CPRD-registered patients), L-TOT users (over incident opioid users), and L-TOT discontinuers (over L-TOT users) were calculated. Annual counts of each group were fitted using segmented negative binomial regression models with an offset considering their corresponding denominators from 2009 to 2019, excluding 2014 due to policy changes in that year.

**Results:** Among 2,839,161 incident opioid users, 11.4% (n=324,877) transitioned into L-TOT users within one year, of which 4.8% (n=15,484) discontinued. Between 2009-2013, rates of L-TOT users significantly declined by 2.6% (incidence rate ratio: 0.974; 95% confidence interval: 0.971 to 0.978) per annum, followed by a significant step change in 2015 (1.026, 1.009 to1.044), and a significant annual increase of 2.4% (1.024, 1.019 to 1.029) from 2015 to 2019, compared to the 2009-2013 trend. The annual rates of L-TOT discontinuers remained stable from 2009 to 2013 (0.987, 0.971 to 1.002), followed by a non-significant step change (0.990, 0.916 to 1.070) in 2015, and a significant decrease in slope by 2.6% (0.974, 0.951 to 0.998) per annum during 2015-2019, relative to 2009-2013.

**Conclusions:** L-TOT has plateaued since 2015, accompanied with an accelerated decrease in discontinuation rates, suggesting ongoing reliance on opioids for CNCP management, despite increased awareness regarding L-TOT associated risks. This is likely due to the limited availability of other effective pharmacological options and non-pharmacological alternatives, and challenges in their accessibility.

**What is already known on this topic:**

- Opioid prescribing for CNCP has increased markedly in the UK over the past two decades. CNCP patients are particularly susceptible to prolonged opioid use, often transitioning to L-TOT due to the complexity of pain management.
- L-TOT significantly increases the risk of opioid-related harms, including dependence, addiction, bone fractures, and opioid-related death.
- Regulatory interventions and awareness of opioid-related harms have led to a decrease in opioid prescribing since 2014. However, research on trends in L-TOT, especially its discontinuation, is lacking in UK primary care. There is an imperative need to investigate if the trends for L-TOT and its discontinuation align with current clinical recommendations.

**What this study adds:**

- Incident opioid prescribing rates declined significantly after 2015, likely due to the impact of policy measures (e.g., the 2014 tramadol reclassification policy) and increasing awareness of opioid-related risks.
- Among incident opioid users, around 1 in 10 transitioned to L-TOT, which showed a decreasing trend followed by a plateau in recent years, suggesting an ongoing reliance on opioids for CNCP management.
- The discontinuation rate of L-TOT remained low at 4.8%, and it significantly declined from 2015 onwards, highlighting challenges in L-TOT discontinuation, such as limited options for other effective medications, and challenges with accessing non-pharmacological alternatives.

## Introduction

Opioid prescribing for chronic non-cancer pain (CNCP) has increased markedly in the United Kingdom (UK) over the past two decades, ^1, 2^ raising particular concerns for these people. Due to the complexity of pain management, CNCP patients are more likely to transition to long-term opioid therapy (L-TOT), which in turn exposes them to increased risks of dependency, addiction, bone fractures and opioid-related death. ^3–5^ In response, several national prescribing guidelines ^6–9^ in the UK have recommended reducing opioid prescribing, thereby to mitigate L-TOT and its associated harms. In fact, prior to the implementation of these guidelines, growing awareness among healthcare professionals about the adverse effects of opioids, especially in their long-term use, had already led to a levelling-off in opioid prescribing since 2014. ^3, 10^ Regulatory interventions, such as the 2014 reclassification of tramadol as a Schedule III controlled drug, ^11^ which imposed stricter prescribing and dispensing controls of tramadol, and the earlier 2013 national Controlled Drugs (Supervision of Management and Use) Regulations, ^12^ which aimed at improving the monitoring and use of controlled drugs such as opioids, may have further promoted this trend. These changes have marked an important turning point, calling for further examination of how the trend of opioid prescribing has changed over time. Such an analysis will help assess whether current practices align with recommendations, particularly for L-TOT and its subsequent discontinuation in UK primary care, where opioids are predominantly prescribed. ^13^

Current evidence on the trends of L-TOT primarily came from the United States (US) and focused on the general population. ^14^ In the UK, although some studies have reported such data, ^2, 15^ they were limited to specific conditions, such as musculoskeletal pain or broader non-cancer pain (not CNCP), and were restricted to earlier periods, up to 2013. Therefore, there is a gap in understanding the trends of L-TOT beyond this period, especially within the CNCP population, who often require long-term pain management and face greater risks from L-TOT than the general population. ^16, 17^ L-TOT discontinuation is essential in reducing potential opioid-related harms. Understanding its level and trend over a recent period will help to identify how well policy and clinical practice have affected the management of L-TOT in UK primary care. However, current evidence regarding such information is lacking, and the reported discontinuation rates derived from US-based studies ranging widely from ∼7% to 72%, ^18–23^ depending on the definitions and methodologies used. These findings may not be directly applicable to the UK context due to the differences in healthcare systems, prescribing practices, and patient demographics, which highlight the need for research in this area.

This study aimed to address these gaps by examining patterns and trends of opioid use among people with CNCP in UK primary care, with a special focus on L-TOT and its discontinuation. By analysing these trends, this study sought to provide insights to inform future policies and clinical practice, ultimately promoting safer opioid prescribing and improving the management of CNCP in UK primary care.

## Methods

### Study design and population

This retrospective cohort study used data from the Clinical Practice Research Datalink (CPRD) Aurum, a database of anonymised UK primary care electronic health records, ^24^ covering 20% of the national population (approximately 13 million active registrants). We included people aged ≥18 years on the index date (the first opioid prescription following a 12-month opioid-free period) with a diagnosis of CNCP (code lists are provided in Supplementary Table S1), and received ≥1 opioid prescription between 1^st^ January 2009 and 31^st^ December 2019. CNCP diagnosis could occur up to 6 months after or any time before the index date. Patients with a cancer diagnosis, except for non-melanoma skin cancer, in the previous ten years were excluded due to different drug utilisation patterns and different guidelines for opioid prescribing. To be eligible, patients were required to have been registered with the CPRD practice for at least 12 months, and have available linked data for both the Index of Multiple Deprivation (IMD) 2019 and the Hospital Episode Statistics Admitted Patient Care database (HES APC).

### Opioid measures

Incident opioid users were defined as having no opioid prescription record for ≥12 months prior to the date of the next prescription. L-TOT users were defined as receiving ≥ 3 opioid prescriptions within a 90-day period, or a sum of opioid supply days lasting ≥ 90 days within the first year of follow-up, excluding the initial 30 days. ^2^ L-TOT discontinuers were defined as no opioid use for ≥180 days following an episode of L-TOT. The study cohort identification process is shown in Supplementary Figure S1.

All nonparental opioid analgesics (code list is provided in Supplementary Table S2) were included, except for methadone and sublingual buprenorphine, as their primary indication is for opioid addiction rather than pain management. Opioid drug exposure data were processed using the logic of a previously published drug preparation algorithm. ^2, 25^ Details of the relevant decision process are provided in Supplementary Figure S2. Opioid doses were converted to oral morphine milligram equivalents (MME) using standardised conversion factors. ^26^

### Covariates

Baseline patient characteristics (i.e., age, sex) were collected on the index date. Ethnicity data was obtained through the linkage with HES APC data. ^27^ Regions were classified as North England (North East, North West, Yorkshire & the Humber), Midland (East Midlands, West Midlands), South England (South West, South Central, South East Coast, East of England), and London. Socioeconomic status was measured using the IMD 2019 ^28^ and categorised into quintiles (Q1-least deprived, Q5-most deprived).

Comorbidities were measured by the Charlson comorbidity index (CCI) score and categorised into three grades: mild (0-2); moderate (3-4); and severe (≥5). ^29^ The presence of other conditions, including substance use disorders (SUDs), alcohol dependence, anxiety, depression, schizophrenia, osteoarthritis, rheumatoid arthritis, and epilepsy were identified within five years prior to the index date. We also identified concurrent drug use including benzodiazepines, gabapentinoids, antidepressants, Z-drugs, muscle relaxants or non-steroidal anti-inflammatory drugs (NSAIDs) within five years prior to the index date.

### Statistical analysis

Descriptive statistics were used to present patient baseline characteristics, including demographics, socioeconomic status, lifestyle (i.e., drinking and smoking status), comorbidities, and concurrent drug use. The yearly rates of incident opioid users, L-TOT users, and L-TOT discontinuers were calculated using below formulae:

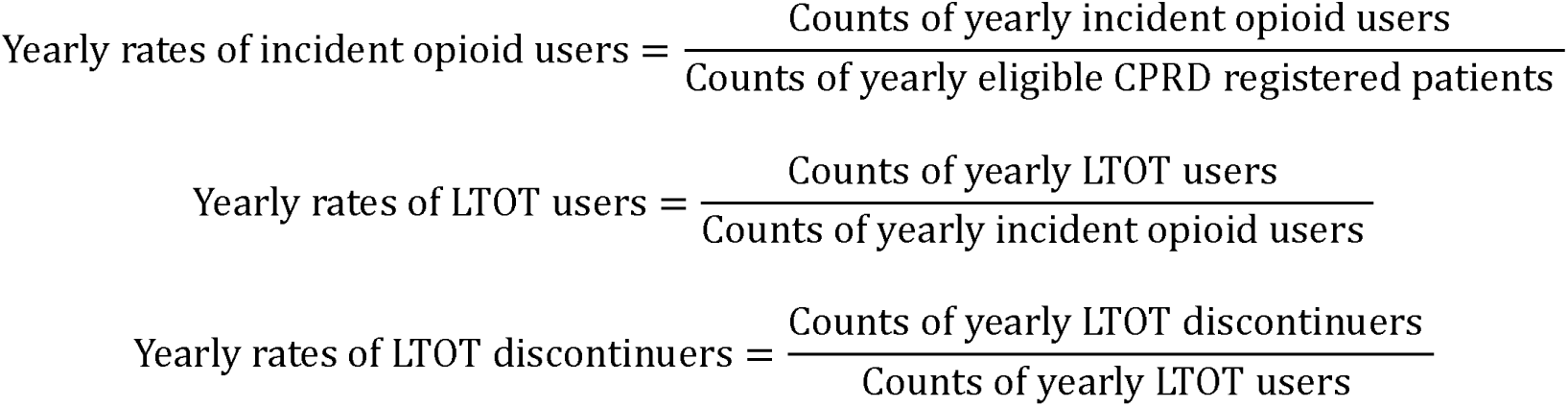

Annual counts of each group (2009-2019) were fitted using segmented negative binomial regression models, with an offset term based on the natural logarithm of their corresponding denominators: CPRD registered patients for incident opioid users, 2014 was selected as the breakpoint based on previous studies ^10, 30^ and structural break analysis (Supplementary Table S3). However, we *a-priori* decided to exclude the 2014 data from our main analyses to avoid introducing potential bias from a partial effect of the tramadol reclassification policy in that year. ^11^

All models included: 1) a year variable indicating the yearly trend from 2009 to 2013 (β1); 2) a binary indicator variable differentiating two time periods (coded ‘0’ for the period of 2009-2013 and ‘1’ for the period of 2015-2019), reflecting the step (level) change (β2) between the observed in 2015 and the projected in 2015 (a linear projection obtained based on the 2009-2013 data); and 3) an interaction term between year and the indicator variable, quantifying the annual slope change (β3) during 2015-2019 compared to the 2009-2013 slope. All data analyses were conducted with Stata/MP 18.0.

### Sensitivity analysis

To assess the robustness of our results, we conducted two sensitivity analyses: 1) including the year 2014. 2) we performed Poisson regression as an alternative to negative binomial regression.

### Patient and public involvement

We have engaged patient groups, including the PRIMER (Primary Care Research in Manchester Engagement Resource, https://sites.manchester.ac.uk/primer/) at the University of Manchester, to assist in developing our dissemination strategy.

## Result

### Patient Characteristics

This study included 2,839,161 incident opioid users between 1^st^ January 2009 and 31^st^ December 2019, of which 11.4% (n=324,877) transitioned to L-TOT users within one year. Among these L-TOT users, 4.8% (n=15,484) subsequently discontinued (Supplementary Figure S3) in the following one year. Table 1 summarised the demographics, comorbidities, concurrent medication use, and opioid dosage and formulation for L-TOT users and L-TOT discontinuers. In both groups, over 78% were white, and females constituted more than 57%. L-TOT discontinuers were generally younger (mean age: 55.58±18.87 years) and had a lower average daily dose (18.80±11.32 MME mg/day). Compared to L-TOT users, they also had higher proportions of anxiety (35.26%), depression (43.46%), lower CCI scores (73.88% had a score of 0-2), lived in London (15.38%), used weak opioids (82.83%) or short-acting opioids (96.76%), and had concurrent use of NSAIDs (63.87%), and benzodiazepines (22.35%).

**Table 1.** Demographic, comorbidity, concurrent drug, and opioid characteristics, among L-TOT users and L-TOT discontinuers between 2009-2019.

|  |  | **L-TOT users (n= 309,393)** | **L-TOT discontinuers (n=15,484)** |
| --- | --- | --- | --- |
| Sex | Female | 177,305 (57.31%) | 8,962 (57.88%) |
|  | Male | 132,088 (42.69%) | 6,522 (42.12%) |
| Age (mean±SD) |  | 58.39±18.69 | 55.58±18.87 |
| Age group | 18-29 years | 22,301 (7.21%) | 1,519 (9.81%) |
|  | 30-39 years | 34,673 (11.21%) | 2,087 (13.48%) |
|  | 40-49 years | 48,275 (15.60%) | 2,574 (16.62%) |
|  | 50-59 years | 52,026 (16.82%) | 2,537 (16.38%) |
|  | 60-69 years | 55,275 (17.87%) | 2,633 (17.00%) |
|  | 70-79 years | 49,767 (16.09%) | 2,308 (14.91%) |
|  | 80-89 years | 36,460 (11.78%) | 1,446 (9.34%) |
|  | ≥90 years | 10,616 (3.43%) | 380 (2.45%) |
| IMD quintile | 1 least deprived | 51,629 (16.69%) | 2,515 (16.24%) |
|  | 2 | 57,984 (18.74%) | 2,828 (18.26%) |
|  | 3 | 57,087 (18.45%) | 2,988 (19.30%) |
|  | 4 | 62,645 (20.25%) | 3,245 (20.96%) |
|  | 5 most deprived | 73,614 (23.79%) | 3,598 (23.24%) |
|  | Missing | 6,434 (2.08%) | 310 (2.00%) |
| Region | North England | 95,012 (30.71%) | 4,345 (28.06%) |
|  | Midland | 62,124 (20.08%) | 3,163 (20.43%) |
|  | South England | 112,703 (36.43%) | 5,594 (36.13%) |
|  | London | 39,554 (12.78%) | 2,382 (15.38%) |
| Ethnicity | White | 252,747 (81.69%) | 12,092 (78.09%) |
|  | Asian | 11,582 (3.74%) | 863 (5.57%) |
|  | Black | 7,678 (2.48%) | 560 (3.62%) |
|  | Mixed | 1,775 (0.57%) | 120 (0.77%) |
|  | Other | 3,745 (1.21%) | 227 (1.47%) |
|  | Unknown | 31,866 (10.30%) | 1,622 (10.48%) |
| Opioid characteristics | Average daily dose (MME mg/day) | 20.16±11.81 | 18.80±11.32 |
|  | <50 MME mg/day | 306,492 (99.06%) | 15,391 (99.40%) |
|  | [50, 90) MME mg/day | 2,646 (0.86%) | 85 (0.55%) |
|  | [90,120) MME mg/day | 182 (0.06%) | 5 (0.03%) |
|  | ≥120 MME mg/day | 73 (0.02%) | 3 (0.02%) |
|  | Weak opioids | 250,465 (80.95%) | 12,826 (82.83%) |
|  | Short-acting opioids | 294,134 (95.07%) | 14,983 (96.76%) |
| Drinking status | Current | 166,491 (53.81%) | 8,241 (53.22%) |
|  | Never | 49,890 (16.13%) | 2,485 (16.05%) |
|  | Former | 3,872 (1.25%) | 179 (1.16%) |
|  | Unknown | 89,140 (28.81%) | 4,579 (29.57%) |
| Smoking status | Current | 66,861 (21.61%) | 3,224 (20.82%) |
|  | Never | 143,377 (46.34%) | 7,448 (48.10%) |
|  | Former | 98,728 (31.91%) | 4,796 (30.97%) |
|  | Unknown | 437 (0.14%) | 16 (0.10%) |
| Comorbidity | RA | 9,686 (3.13%) | 447 (2.89%) |
|  | OA | 31,079 (10.05%) | 1,420 (9.17%) |
|  | Anxiety | 105,145 (33.98%) | 5,459 (35.26%) |
|  | Depression | 133,616 (43.19%) | 6,729 (43.46%) |
|  | Schizophrenia | 7,791 (2.52%) | 333 (2.15%) |
|  | Alcohol dependence | 14,243 (4.60%) | 595 (3.84%) |
|  | Epilepsy | 9,419 (3.04%) | 413 (2.67%) |
|  | Substance Misuse Disorder | 13,934 (4.50%) | 552 (3.56%) |
| CCI | Mild (0-2) | 222,177 (71.81%) | 11,439 (73.88%) |
|  | Moderate (3-4) | 53,199 (17.19%) | 2,511 (16.22%) |
|  | Severe (≥5) | 34,017 (10.99%) | 1,534 (9.91%) |
| Concurrent drug use | Benzodiazepines | 66,976 (21.65%) | 3,461 (22.35%) |
|  | Antidepressants | 144,542 (46.72%) | 6,986 (45.12%) |
|  | Gabapentinoids | 2,440 (0.79%) | 98 (0.63%) |
|  | Muscle relaxants | 3,283 (1.06%) | 169 (1.09%) |
|  | Z drugs | 39,757 (12.85%) | 1,953 (12.61%) |
|  | NSAIDs | 187,018 (60.45%) | 9,889 (63.87%) |

Note: IMD = Index of Multiple Deprivation; MME = Morphine Milligram Equivalents; RA = Rheumatoid Arthritis; OA = Osteoarthritis; CCI = Charlson Comorbidity Index; NSAIDs
= Non-steroidal anti-inflammatory drugs; L-TOT users means those who did not discontinue opioids

### Trends for incident opioid users, L-TOT users and L-TOT discontinuers

Table 2,3 and Figure 1 illustrated the trends of incident opioid users, L-TOT users, and L-TOT discontinuers. Between 2009 and 2013, the yearly rates of incident opioid users showed a flat trend (incidence rate ratio: 1.001, 95% confidence interval: 0.988 to 1.014). Although there was a non-significant step change in 2015 (0.988, 0.928 to 1.052), their trend from 2015 to 2019 showed a significant annual decrease of 8.7% (0.913, 0.897 to 0.930), compared to the slope during 2009-2013.

**Table 2.** The yearly numerator, denominator and rate of incident opioid users, L-TOT users and L-TOT discontinuers from 2009 to 2019.

| Year | No. CPRD registered participants | No. Incident opioid users | No. L-TOT users | No. L-TOT discontinuers | Rate of incident opioid users (%) | Rate of L-TOT users (%) | Rate of L-TOT discontinuers (%) |
| --- | --- | --- | --- | --- | --- | --- | --- |
| 2009 | 1,165,440 | 254,066 | 31,905 | 1,690 | 21.80 | 12.56 | 5.30 |
| 2010 | 1,194,284 | 262,411 | 31,978 | 1,620 | 21.97 | 12.19 | 5.07 |
| 2011 | 1,218,921 | 266,417 | 32,001 | 1,589 | 21.86 | 12.01 | 4.97 |
| 2012 | 1,240,969 | 274,116 | 31,515 | 1,570 | 22.09 | 11.50 | 4.98 |
| 2013 | 1,260,446 | 275,520 | 31,261 | 1,569 | 21.86 | 11.35 | 5.02 |
| 2014 | 1,358,015 | 280,519 | 31,749 | 1,470 | 20.66 | 11.32 | 4.63 |
| 2015 | 1,326,781 | 278,251 | 30,480 | 1,416 | 20.97 | 10.95 | 4.65 |
| 2016 | 1,328,107 | 269,706 | 29,697 | 1,391 | 20.31 | 11.01 | 4.68 |
| 2017 | 1,332,717 | 249,669 | 27,646 | 1,237 | 18.73 | 11.07 | 4.47 |
| 2018 | 1,323,147 | 224,008 | 24,424 | 1,057 | 16.93 | 10.90 | 4.33 |
| 2019 | 1,392,118 | 204,478 | 22,221 | 875 | 14.69 | 10.87 | 3.94 |

**Table 3.** Incidence rate ratios of negative binominal regression models for yearly rates of incident opioid users, L-TOT users and L-TOT discontinuers during 2009-2013 and 2015-2019.

|  | Incident opioid user<br>(n=2,558,642) | L-TOT users<br>(n=293,128) | L-TOT Discontinuers<br>(n=14,014) |
| --- | --- | --- | --- |
| Regression model | IRR, 95%CI | IRR, 95%CI | IRR, 95%CI |
| Time ($\beta_1$) | 1.001 (0.988, 1.014) | 0.974 (0.971, 0.978) * | 0.987 (0.972, 1.003) |
| Indicator ($\beta_2$) | 0.988 (0.928, 1.052) | 1.026 (1.009, 1.044) * | 0.990 (0.916, 1.070) |
| Interaction term ($\beta_3$) | 0.913 (0.897, 0.930) * | 1.024 (1.019, 1.029) * | 0.974 (0.951, 0.998) * |

Note: IRR=Incidence Rate Ratio, \* p value less than 0.05 was regarded as significantly different

For L-TOT users, during 2009-2013, the yearly rates exhibited a significant decline of 2.6% (0.974, 0.971 to 0.978) per annum, followed by a statistically significant step change in 2015 (1.026, 1.009 to 1.044), and an annual increase of 2.4% thereafter from 2015 to 2019 (1.024, 1.019 to 1.029), relative to the trend from 2009 to 2013. In summary, the trend for L-TOT users decreased initially but plateaued in recent years.

The annual rates of L-TOT discontinuation showed a relatively stable trend from 2009 to 2013, with a non-significant annual decrease of 1.3% (0.987, 0.971 to 1.002), followed by a non-significant step change in 2015. This downward trend continued but became significant during 2015-2019, with a decrease of 2.6% per annum (0.974, 0.951 to 0.998) compared to the slope between 2009 and 2013. In summary, L-TOT discontinuation began to decrease significantly only from 2015 onwards.

### Sensitivity analysis

The sensitivity analysis including the year 2014 data showed that the pre-post 2014 trends of yearly rates for incident opioid users and L-TOT users maintained consistent with the main analyses (Supplementary Table S4). However, the annual L-TOT discontinuation rates in the pre-2014 trend became significant (0.980, 0.969 to 0.992), whereas the post-2014 slope change turned to be non-significant (0.981, 0.960 to 1.003). However, despite these changes in significance, the overall trend indicated a steady decline in discontinuation rates throughout the study period from 2009 to 2019. Results from Poisson regressions were consistent with those obtained from the negative binominal regressions (Supplementary Table S5).

## Discussion

In this population-based retrospective cohort study, we analysed the trends of incident opioid users, L-TOT users and L-TOT discontinuers from 2009-2019, using 2014 as the change point. Between 2009-2013, rates of incident opioid users remained stable but showed a significant annual decline from 2015 onward, highlighting the effectiveness of the 2014 tramadol reclassification ^11^ and other regulatory interventions ^12^ in reducing opioid initiations. However, the trend for L-TOT users plateaued since 2015, despite a previously downward trend between 2009-2013. This stabilisation can be attributed, at least partly, to the sharp reduction in incident opioid users, which resulted in a smaller pool of patients eligible for transitioning to L-TOT. At the same time, this shift highlights a persistent reliance on opioids for managing CNCP in certain patient groups, for whom, L-TOT may still be perceived as essential despite the known risks and ongoing policy interventions.

Regarding L-TOT discontinuation, the overall rate remained low at 4.8%, indicating substantial barriers to discontinuation and many patients being exposed to the associated risks. Factors contributing to this may include physiological dependence, fear of withdrawal symptoms, limited access to effective non-opioid pain management strategies, and clinicians’ hesitancy to initiate reductions. ^31–33^ Over time, L-TOT discontinuation rates steadily declined, with a marked drop after 2015. This decline likely reflects changes in both the numerator (fewer patients discontinuing L-TOT) and the denominator (a changing L-TOT user population). With fewer new patients initiating opioids, the remaining L-TOT population may increasingly consist of individuals with more complex, treatment-resistant conditions. These patients are less likely to discontinue opioids due to their greater reliance on L-TOT for managing severe or persistent pain. Additionally, the impact of regulatory measures and policies may have disproportionately affected new opioid initiations rather than existing L-TOT users, contributing to the observed trends.

### Comparison with other studies

Our study identified an 11.4% rate of L-TOT within the CNCP population, which is comparable to the 14.6% prevalence reported in prior research using CPRD GOLD data among non-cancer pain patients from 2016 to 2021. ^2^ Both studies applied the same definition of L-TOT; however, the slightly lower prevalence in our findings may be explained by the differences in the code lists used to define pain conditions.

In terms of trends, our findings regarding the significant decrease in L-TOT rates from 2009 to 2013, align with an observational study using CPRD GOLD data. ^15^ That study reported a stabilisation in annual long-term opioid incidence (defined as receiving ≥2 opioid prescriptions within 90 days) among patients with musculoskeletal pain from 2009 to 2011, followed by a slight decline by 2013. While there are differences in study populations and definitions of L-TOT, the trends are generally comparable.

In contrast, data from the NHS Business Services Authority (BSA)’s medication safety indicators dashboard showed a 54% increase in the number of individuals using oral or transdermal opioids for longer than three months in England between 2015 and 2021. ^34^ This finding was contradictory to our study, which observed a plateau in L-TOT rates during the similar timeframe. The discrepancies may be explained by differences in the definition of L-TOT and study design, i.e., the NHS BSA data does not specify its study population and likely includes a broader demographic beyond CNCP patients. However, despite these differences, both our study and the NHS BSA data indicated that L-TOT use has not significantly declined in recent years. This persistent trend again, highlighted the ongoing challenges in stopping long-term opioid prescribing.

Focusing on L-TOT discontinuation, current evidence on its prevalence is predominantly derived from US-based studies, with reported rates varying widely from ∼7% to 72%, often among veterans with CNCP or populations receiving high-dose chronic opioid therapy (>90 MME mg/day). ^18–23^ Our study identified a lower L-TOT discontinuation rate of 4.8% in the CNCP population, who were prescribed with lower daily doses (mean ∼18 MME mg/day). This conservative opioid prescribing pattern reflected a cautious approach taken by general practitioners (GPs) in the UK, aligning with current clinical guidelines, ^35^ which recommend minimising opioid doses not to exceed 50 MME mg/day whenever possible. However, it is important to note that differences in study populations, settings, and definitions of both L-TOT and discontinuation limit the direct comparability of our findings with those from the US. Nevertheless, this low discontinuation rate emphasised two challenges: first, an ongoing reliance on opioids for managing CNCP in UK primary care despite guidelines ^8, 36^ discouraging them as the first-line treatment; second, the significant difficulties mentioned above in stopping L-TOT among this population.

To date, no UK-based studies have reported the trends of L-TOT discontinuation. The only evidence came from a US study, ^18^ which reported an increase in annual L-TOT discontinuation rates from 5.7% in 2012 to 8.5% in 2017 among Medicare beneficiaries with non-cancer pain. While this upward trend contrasted with our observed declining-to-flat L-TOT discontinuation rates, these results were not comparable due to variations in definitions, study populations, healthcare systems, and pain conditions. Nevertheless, both studies highlighted the reliance on L-TOT, as noted by the US authors in their paper, that the majority of patients remained on L-TOT every year.

Unlike the US, where opioid monitoring programmes ^37^ and proactive opioid taper decision tools ^38^ are often available, the UK currently lacks such tailored interventions, development of evidence-based, patient-centred approaches, along with alternative pain management strategies that align with current clinical guidelines. Such efforts are critical to overcoming the barriers to L-TOT discontinuation and reducing opioid reliance in the management of CNCP.

### Strengths and limitations of this study

This study has several strengths. First, to our knowledge, this is the first study using CPRD Aurum data to analyse trends in opioid use, particularly focusing on the L-TOT and its subsequent discontinuation among people with CNCP in UK primary care. Previous research has not addressed the trends in L-TOT discontinuation, making this study an important addition to the literature. Second, CPRD Aurum database provided comprehensive, high-quality data ^39^, and the large and nationally representative sample ensured the robustness and generalisability of our findings to the UK population. Finally, the code lists used in this study were developed and validated by a research team member (CM), which ensured the methodological rigor.

Some limitations need to be acknowledged. First, while the CPRD Aurum database provides comprehensive information on prescribed opioids, it does not capture over-the-counter opioids such as codeine and dihydrocodeine, which may potentially underestimate total opioid utilisation in the study population. Second, the availability of prescription records does not guarantee that patients actually used the medications as prescribed, as we cannot observe adherence. This limitation is inherent to the use of CPRD data and the reliance on prescription records. Third, the findings may not be fully generalisable to other healthcare settings or populations with different prescribing practices or guidelines. Fourth, although the analysis focused on trends before and after 2014, these trends may have been influenced by other concurrent national policies on analgesic prescribing and the exclusion of this year may limit the ability to fully assess the impact of policy change on prescribing practices. Fifth, the regression analysis did not account for all potential confounding factors (e.g., pain severity, marital status, healthcare insurance, etc.), which would provide a more complete picture of opioid use and its effects. Lastly, we focused on discontinuation rather than dose reductions. Many L-TOT users may reduce their doses without fully stopping, which could potentially underestimate efforts to reduce opioid prescribing in primary care.

### Clinical implications

This study has important clinical implications for L-TOT discontinuation and the management of CNCP in UK primary care. Incident opioid prescribing for CNCP has decreased in recent years, reflecting the impact of policies and growing awareness of opioid-related risks. Although these efforts have contributed to a reduction in L-TOT use, achieving a consistent decline remains challenging. It is concerning that 11.4% of individuals still transitioned to L-TOT, highlighting the difficulties faced by clinicians and patients in stopping or reducing opioid use. ^40^ These challenges include the lack of safe and effective pharmacological alternatives, the complex nature of chronic pain, and barriers that prevent patients from accessing or engaging in non-pharmacological pain treatments that they find acceptable, such as physical therapy, exercise, or cognitive-behavioural therapy. Often these treatments and therapies need adapting for marginalised groups.

The decline in L-TOT discontinuation rates further suggested that patients are struggling with stopping L-TOT. Alongside carefully assessing the necessity of opioid prescriptions, and limiting duration of prescriptions, our results suggested GPs should aim to identify patients who are likely to succeed in discontinuation. To support GPs in this, clear discontinuation protocols need to be developed at a local and/or national level. ^9^ Furthermore, adopting a holistic pain management approach, addressing both physical, social and psychological aspects of CNCP could complement these efforts to improve patient outcomes and promote safer opioid use. ^41^

### Conclusions

In conclusion, this study provided valuable insights into the trends of L-TOT use and its discontinuation for CNCP in UK primary care. The decreasing-to-flat trend of L-TOT users and the consistent declining trend of L-TOT discontinuers suggested an ongoing reliance on opioids for CNCP management despite policy impacts and growing awareness of L-TOT associated risks, which likely due to the lack of safe and effective pharmacological alternatives, the complexity of chronic pain management, and factors making it difficult for patients to access or engage in acceptable non-pharmacological pain treatments. These findings highlighted the need for sustained efforts to promote safer prescribing and discontinuation practices. Future research should identify key factors that contribute to successful L-TOT discontinuation, barriers which prevent discontinuation, and evaluate the effectiveness of opioid reduction strategies.

## Ethics statements

### Ethical approval

This study was approved by the CPRD’s Independent Scientific Advisory Committee (protocol number: 23_002909, available at https://www.cprd.com/approved-studies/predictors-discontinuation-or-reduction-long-term-opioid-therapy-and-its) and was reported according to the Strengthening the Reporting of Observational Studies in Epidemiology (STROBE) Statement (Supplementary Table S6). ^42^

## Data availability

This study used anonymised individual-level data from the CPRD Aurum. The data cannot be shared publicly due to CPRD regulations and ethical reasons. Other researchers can use the CPRD data in a secure environment by submitting a research protocol to the CPRD Independent Scientific Advisory Committee. Details of the application process and conditions of access are provided by the CPRD at https://www.cprd.com/Data-access.

## Acknowledgments

We gratefully acknowledge Professor Daren Ashcroft and the NIHR Greater Manchester Patient Safety Research Collaboration, for covering the data access costs. We also thank all contributing patients and general practitioners for making anonymised data available for research.

## Footnotes

### Contributors

QC, EK, TA, CG, YTH conceived and designed the study. EK, TA, CG and YTH supervised the conduct of the study. CM reviewed the code lists. QC conducted the data analysis and drafted the initial version of the manuscript. QC, EK, TA, CG, YTH contributed to the interpretation of the findings. All authors have reviewed the manuscript and contributed to revisions. QC is the guarantor. The corresponding author (QC) attests that all listed authors meet authorship criteria and that no others meeting the criteria have been omitted.

### Transparency

The lead author (QC) confirms that the manuscript is an honest, accurate, and transparent account of the study, with no important omissions or unexplained discrepancies from the planned study.

### Dissemination to participants and related patient/public communities

As this study used anonymised CPRD data, direct dissemination to individuals is not possible. The University of Manchester networks will further disseminate the results, as the study forms a significant part of QC’s PhD.

---

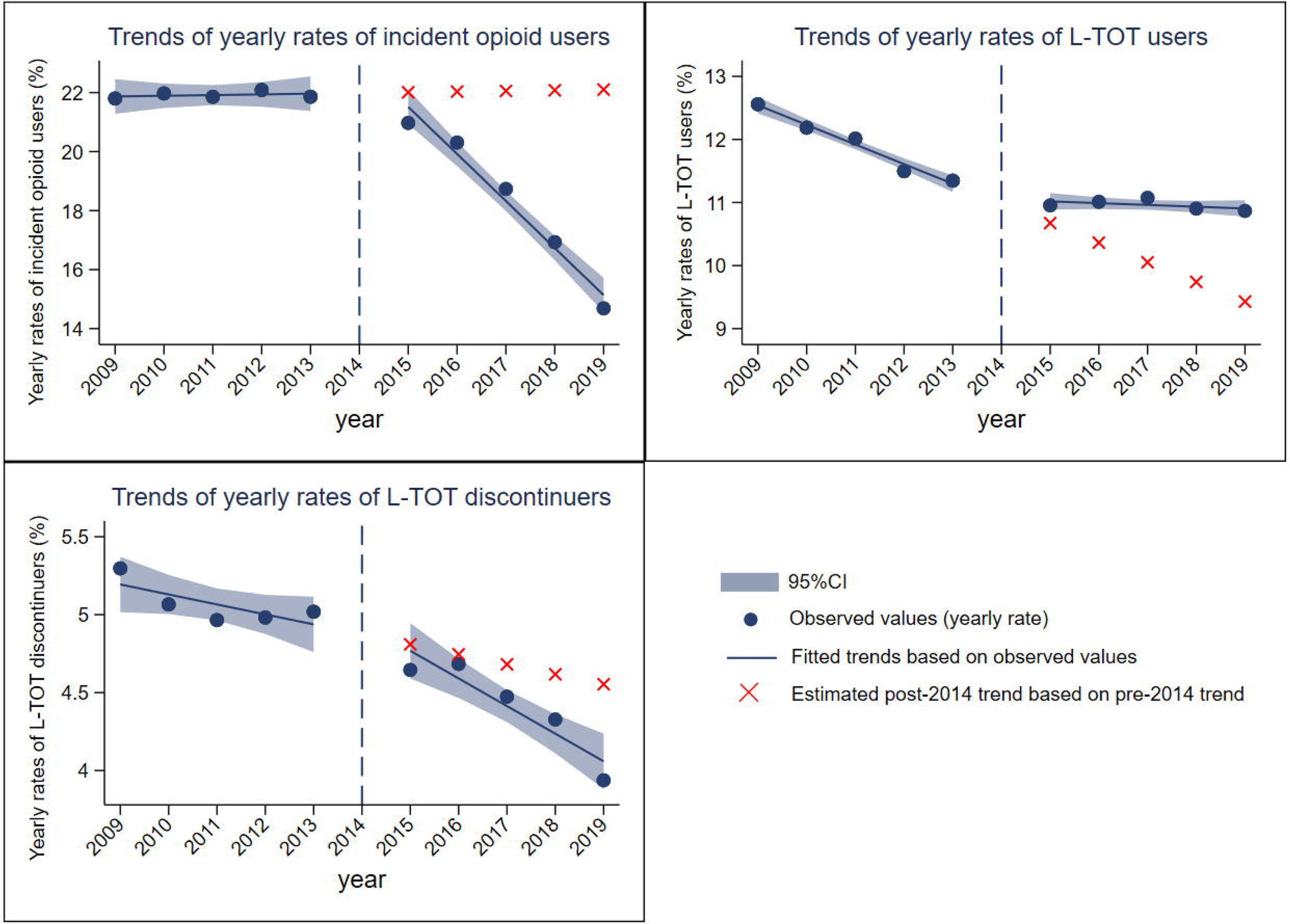

